# LUCAS: A highly accurate yet simple risk calculator that predicts survival of COVID-19 patients using rapid routine tests

**DOI:** 10.1101/2021.04.27.21256196

**Authors:** Surajit Ray, Andrew Swift, Joseph W Fanstone, Abhirup Banerjee, Michail Mamalakis, Bart Vorselaars, Louise S Mackenzie, Simonne Weeks

## Abstract

**Background:** There is an urgent need to develop a simplified risk tool that enables rapid triaging of SARS CoV-2 positive patients during hospital admission, which complements current practice. Many predictive tools developed to date are complex, rely on multiple blood results and past medical history, do not include chest X ray results and rely on Artificial Intelligence rather than simplified algorithms. Our aim was to develop a simplified risk-tool based on five parameters and CXR image data that predicts the 60-day survival of adult SARS CoV-2 positive patients at hospital admission.

**Methods:** We analysed the NCCID database of patient blood variables and CXR images from 19 hospitals across the UK contributed clinical data on SARS CoV-2 positive patients using multivariable logistic regression. The initial dataset was non-randomly split between development and internal validation dataset with 1434 and 310 SARS CoV-2 positive patients, respectively. External validation of final model conducted on 741 Accident and Emergency admissions with suspected SARS CoV-2 infection from a separate NHS Trust which was not part of the initial NCCID data set.

**Findings:** The LUCAS mortality score included five strongest predictors (lymphocyte count, urea, CRP, age, sex), which are available at any point of care with rapid turnaround of results. Our simple multivariable logistic model showed high discrimination for fatal outcome with the AUC-ROC in development cohort 0.765 (95% confidence interval (CI): 0.738 - 0.790), in internal validation cohort 0.744 (CI: 0.673 - 0.808), and in external validation cohort 0.752 (CI: 0.713 - 0.787). The discriminatory power of LUCAS mortality score was increased slightly when including the CXR image data (for normal versus abnormal): internal validation AUC-ROC 0.770 (CI: 0.695 - 0.836) and external validation AUC-ROC 0.791 (CI: 0.746 - 0.833). The discriminatory power of LUCAS and LUCAS + CXR performed in the upper quartile of pre-existing risk stratification scores with the added advantage of using only 5 predictors.

**Interpretation:** This simplified prognostic tool derived from objective parameters can be used to obtain valid predictions of mortality in patients within 60 days SARS CoV-2 RT-PCR results. This free-to-use simplified tool can be used to assist the triage of patients into low, moderate, high or very high risk of fatality and is available at https://mdscore.net/.

**Summary box:** *What is already known on this topic?:* Clinical prediction models such as NEWS2 is currently used in practice as mortality risk assessment. In a rapid response to support COVID-19 patient assessment and resource management, published risk tools and models have been found to have a high risk of bias and therefore cannot be translated into clinical practice.

*What this study adds?:* A newly developed and validated risk tool (LUCAS) based on rapid and routine blood tests predicts the mortality of patients infected with SARS-CoV-2 virus. This prediction model has both high and robust predictive power and has been tested on an external set of patients and therefore can be used to effectively triage patients when resources are limited. In addition, LUCAS can be used with chest imaging information and NEWS2 score.

## Introduction

The UK National Health Service (NHS) has experienced unprecedented pressure due to the recurrent surges of coronavirus disease 2019 (COVID-19) cases caused by the SARS CoV-2 virus. To date more than two million confirmed cases in the UK have forced healthcare professionals to face complex decisions on how to effectively triage patients on admission who may need acute care. Under these strained conditions there is an urgent need to develop a simplified risk tool that enables rapid triaging of SARS CoV-2 positive patients, which complements predictive models commonly used in practice. Any new model must support the healthcare professional’s experiential knowledge and clinical reasoning [1] in both patient care management and allocation of limited healthcare resources.

Predictive models such as the National Early Warning Score 2 (NEWS2) is widely used in practice to observe and identify deteriorating patients [2]. Although, favourable amongst healthcare professionals [3], this predictive tool has caused an increased false trigger rates for SARS CoV-2 positive patients [4]. Another familiar tool in practice in the UK, is the Acute Physiology And Chronic Health Evaluation II (APACHE II) which is a validated intensive care unit (ICU) scoring tool used to estimate ICU mortality, which has underestimated the mortality risk in SARS CoV-2 positive patients [5]. Several published models aimed to meet the clinician’s need to stratify high-risk SARS CoV-2 positive patients but are yet to be widely clinically implemented and had poor clinician feedback [6]. Here we aim to avoid ‘clinical resistance’ by implementing a new prognostic model by developing and validating a new tool that extends on the widely used NEWS2 model [7]. This model combines clinical data, routine laboratory tests and chest X-ray (CXR) to improve risk stratification and clinical intervention for SARS CoV-2 infection. Studies have shown CXR results add value in the triage of patients who are SARS CoV-2 positive [8]; however, the incremental prognostic value of CXR [9] in addition to clinical data and routine laboratory tests remains to be consolidated in practice.

Using statistical methods used previously [10], we used well-defined predictors and outcomes to limit model overfitting. The model’s development and reporting adhered to the TRIPOD (transparent reporting of a multivariable prediction model for individual prediction or diagnosis) guidelines [11] to publish a simple, user-friendly scoring system for adult patients admitted to hospital with SARS CoV-2 infection that will lead to an improved integration of the best evidence into clinical care pathways that will assist clinicians in risk, patient and resource management. The aim of this study was to develop a simple objective tool for risk stratification in SARS CoV-2 infection that would easily support established triage practice in the hospital setting that integrates results from rapid and routine clinical, laboratory and CXR image data.

## Methods

### Sources of data

A prospective cohort study was conducted with The National COVID-19 Chest Imaging Database (NCCID) [12] that collated clinical data from secondary and tertiary NHS Trusts across the UK with the aim to support SARS CoV-2 care pathways (**Supplementary Table 1**). NHS staff submitted data for patients suspected of SARS CoV-2 with a RT-PCR test that was positive or negative. The centralized data warehouse stored de-identified hospital admission’s clinical data including between 23^rd^ January 2020 and 7^th^ December 2020, which was used for both the development and validation of the mortality risk tool.

**Table 1.** Comparison of Development, Internal and External Dataset Characteristics

| Characteristics | Development<br>(n = 1434) | Internal Validation<br>(n = 310) | External Validation<br>(n = 741) |
| --- | --- | --- | --- |
| Data collection dates | 23/01/20 - 30/04/20 | 01/05/20 - 07/12/2020 | 01/03/20 - 21/08/20 |
| Study Design | Prospective cohort |  | Retrospective cohort |
| Setting | Secondary care hospital admissions from 19 sites across the UK |  | Secondary care hospital A&E admissions from 1 UK site |
| Inclusion criteria | All adult patients with a positive SARS CoV-2 status (derived from cumulative RT-PCR test results) and data at the point of hospital admission. |  |  |
| Outcome | In-hospital mortality within 60 days of positive RT-PCR test |  |  |
| Mortality *n* (%) | 466 (33) | 58 (18) | 326 (32) |
| **Demographics** |  |  |  |
| Ethnicity: White British, Irish or any other background *n* (%) | 498 (35) | 90 (29) | 599 (81) |
| Ethnicity: Mixed white and Caribbean, Asian or any other background *n* (%) | 6 (0) | 0 (0) | 8 (1) |
| Ethnicity: Asian or Asian British background *n* (%) | 84 (6) | 10 (3) | 35 (5) |
| Ethnicity: Black or Black British background *n* (%) | 78 (5) | 2 (1) | 45 (6) |
| Ethnicity: any other ethnicity *n* (%) | 99 (7) | 4 (1) | 14 (2) |
| Ethnicity not stated or reported *n* (%) | 669 (47) | 204 (66) | 40 (5) |
| Median age (IQR), [years] | 73 (48-98) | 74 (52-96) | 76 (53-99) |
| Men *n* (%) | 902 (63) | 175 (57) | 422 (57) |
| **Median clinical data on admission** |  |  |  |
| Duration of symptoms (IQR) [days] | 5 (7) | 3 (6) | - |
| Respiratory rate (IQR) [breaths/min] | 21 (7) | 20 (6) | - |
| Heart rate (IQR) [beats/min] | 90 (25.0) | 89 (26.2) | - |
| PaO<sub>2</sub> (IQR) [% room air] | 93 (84.3)* | 95 (6) | - |
| FiO<sub>2</sub> (IQR) [oxygen flow rate if on O<sub>2</sub>] | 21 (26) | 4 (20.79) | - |
| NEWS2 score (IQR) | 4 (4)* | 3 (5.0)* | - |
| Diastolic BP (IQR) [mmHg] | 74 (18.0) | 74 (17.5) | - |
| Systolic BP (IQR) [mmHg] | 130 (32.0) | 131 (35.0) | - |
| Temperature (IQR) [°C] | 40 (15.0) | 38 (14.0) | - |
| **Median laboratory data on admission** |  |  |  |
| WCC (IQR), [x 10<sup>9</sup>/L] | 7.2 (5.1) | 8.0 (5.4) | 8.1 (5.8) |
| Lymphocytes (IQR), [x 10<sup>9</sup>/L] | 0.9 (0.64) | 1(0.74) | 0.9 (0.8) |
| Platelets (IQR), [x 10<sup>9</sup>/L] | 212 (125.0) | 229 (134.2) | 220 (121.0) |
| D-Dimer (IQR), [µg/L] | - | - | 1118 (1585)* |
| Fibrinogen (IQR), [g/L] | - | - | 5.6 (2.1) |
| CRP (IQR), (mg/L) | 85 (121.5) | 46 (98.2) | 62 (101.0) |
| Ferritin (IQR),# | 541 (230.5)* | 601 (188)* | 483 (697)* |
| Urea (IQR),[mmol/L] | 6.8 (6.0) | 6.5 (5.3) | 7.2 (5.5) |
| Creatinine (IQR), [µmol/L] | 86 (57.0) | 85 (49.0) | 88 (49.5) |
| Troponin I (IQR), [ng/L] | 12 (42.5) | 15 (172) | - |
| Troponin T(IQR), [ng/L] | 22 (40.5)* | 17 (27)* | - |
| Chest X-ray data |  |  |  |
| CXR (abnormal) *n* (%) | 1006 (70) | 172 (55) | 361(49) |
| **Past Medical History** |  |  |  |
| Hypertension *n* (%) | 603 (42) | 144 (46) | - |
| CVS disease *n* (%) | 303 (21) | 66 (21) | - |
| Diabetes mellitus type II *n* (%) | 390 (27) | 92 (30) | - |
| Lung disease *n* (%) | 333 (23) | 79 (26) | - |
| Chronic kidney disease *n* (%) | 279 (20) | 54 (17) | - |
| Current ACEi use *n* (%) | 193 (14) | 53 (17) | - |
| Current Angiotension receptor blocker use *n* (%) | 116 (8) | 24 (8) | - |
| Current NSAID use *n* (%) | 130 (9) | 35 (11) | - |
| **Risk Factors** |  |  |  |
| Smokers *n* (%) | 303 (21) | 82 (27) | - |
| Median pack year history (IQR) | 0 (26) | 15 (40) | - |
| **Deterioration** |  |  |  |
| ITU admission n (%) | 181 (13) | 38 (12) | - |
| Median APACHE score (IQR) | 16 (9.25) | 12.5 (9.75) | - |
| Intubation n (%) | 143 (10)* | 18 (6)* | - |

RT-PCR: reverse transcription polymerase chain reaction; A&E: Accident and Emergency; WCC: White Cell Count; CRP: C-reactive protein
\* indicates missing data values over 40 %;

To evaluate the tool’s generalized performance, a second dataset was used for external validation. The data extracted from the Laboratory Information Management System (LIMS) came from a large NHS Foundation Trust hospital in north-east England UK, which had not participated in the NCCID initiative. Patients who were admitted with suspected SARS CoV-2 infection at the Accident and Emergency (A&E) Department between 1^st^ March 2020 and 21^st^ August 2020 were included. This population was representative of the adult general population and therefore patients aged 16 years and younger were excluded, as the laboratory data thresholds vary compared to adults.

### Participants

Adult patients defined by positive RT-PCR for SARS CoV-2 virus were included from all data sets (development, internal and external validation) for analysis. In order to mitigate the incomplete penetrance of testing during initial stages of the pandemic and limited sensitivity of the RT-PCR swab test, NCCID provided an overall SARS CoV-2 status that was derived from cumulative RT-PCR tests. This approach was also applied to identify SARS CoV-2 positive patients in the external validation data set.

To evaluate the tool’s predictive performance, the NCCID dataset was non-randomly split with the admissions before 30^th^ April 2020 that made up the development cohort (n = 1434) and admission on or after 1^st^ May 2020 that made up the internal validation cohort (n = 310) (**Figure 1**). The external validation cohort (n = 741) was filtered by interaction with A&E dates and mortality dates on all SARS CoV-2 positive patients. This approach was rationalised since patients were more comparable on latent variables such as co-morbid illnesses that were not explicitly captured during the LIMS data extraction of laboratory results. In addition, the filtering on admitted patients increased the density of laboratory results across all the candidate predictors, which made the model less reliant on imputation.

**Figure 1.**
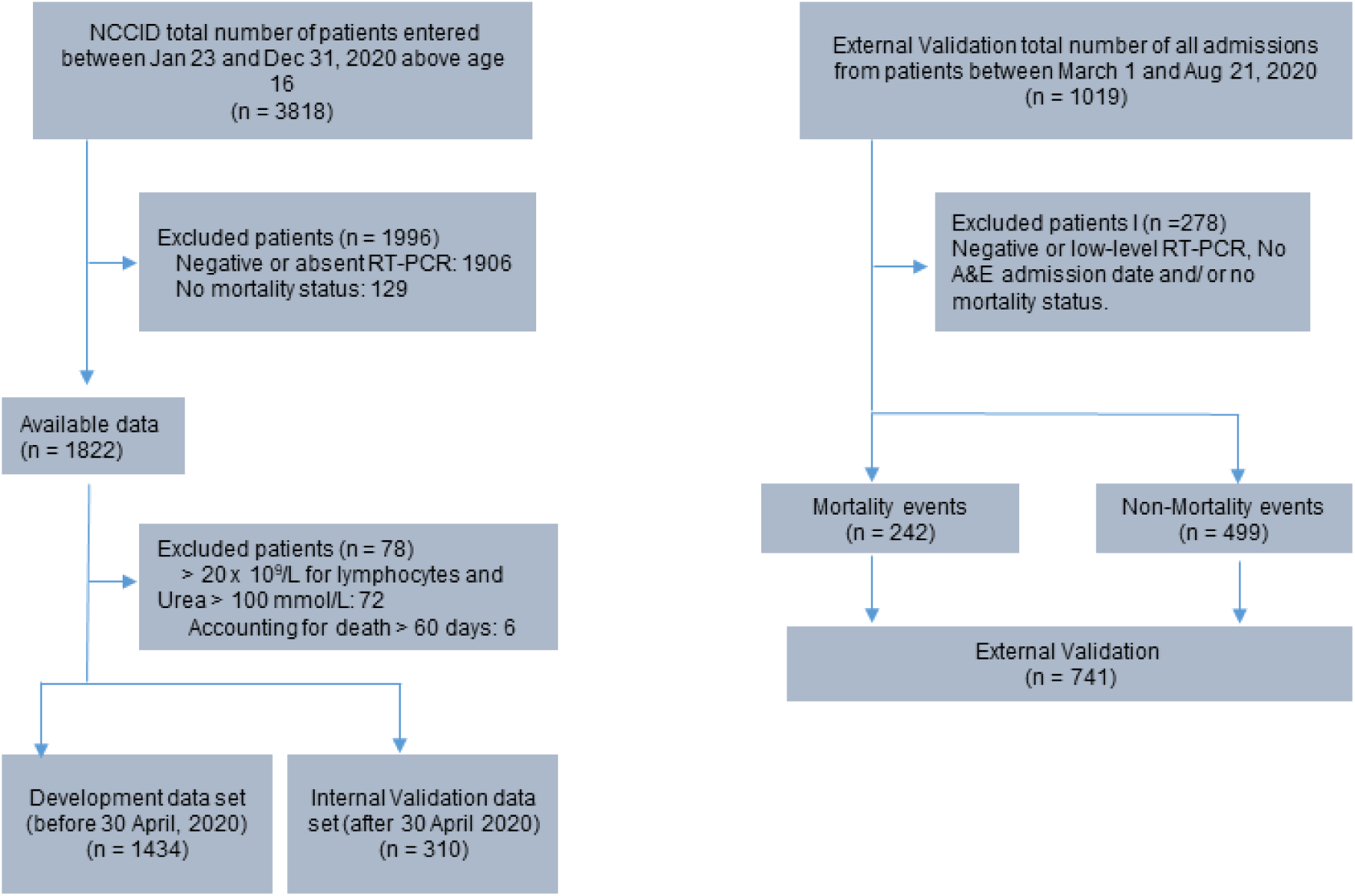
Patient flowchart with inclusion and exclusion criteria that established the development data set as well as internal and external validation data sets.

### Outcome

The primary outcome of interest was in-hospital mortality within 60-days of a positive SARS CoV-2 RT-PCR test. To identify this outcome, the date of death was followed up and entered by hospital staff as part of the NCCID initiative.

### Predictors

The NCCID dataset collected 38 clinical data points for each patient that served as candidate predictors. These were categorised into demographics, risk factors, past medical history (PMH), medications, clinical observations, chest imaging data, and laboratory parameters measured on admission. Chest X-ray (CXR) reports from the NCCID model development dataset and the external dataset were dichotomised as normal versus abnormal. All laboratory tests were measured before SARS CoV-2 RT-PCR test and mortality date, which ensured these potential predictors were blinded to the outcome. The SARS CoV-2 swab and RT-PCR results established the final COVID-19 status. In the event of deterioration, data points such as the NEWS2 score, Acute Physiology and Chronic Health Evaluation (APACHE) score, Intensive Therapy Unit (ITU) admission, intubation and mortality date were also recorded. Since the focus of the study was to identify predictors that were available at the point of admission, the ITU admission and intubation data were excluded in the final analysis.

From this extensive list, only the demographics, clinical observations, and laboratory parameters recorded at the time of admission for SARS CoV-2 positive patients were included in the analysis as key predictors. Previous medical history, smoking status and current pharmaceutical interventions were not included in the analysis. This rationale was based on consultations with biomedical scientists and clinicians working in A&E departments to determine which rapid and routine laboratory tests would be clinically relevant candidate predictors, as well as previously published literature on prognostic factors associated with SARS CoV-2 infection for the model development [13].

To account for the risk of spurious selection or exclusion of important predictors in the model development, a minimum sample size of 380 was determined based on 10 outcome events per variable. Based on previous studies, a minimum of 100 mortality events and 100 non-mortality events was required for external validation sample size [14].

### Statistical Analysis

Reliability of data was ensured by excluding patients from the development and internal validation datasets if information was missing on key characteristics, such as and including RT-PCR SARS CoV-2 positive results, admission and mortality dates outside the 60-day prediction interval (**Figure 2**). Predictors with more than 40% missing values were also excluded from the modelling process (**Supplementary Figure 1**). Missing data from the remaining predictors were handled using Multiple Imputation by Chained Equations (MICE) [15] and ten different imputed data sets were combined using the Rubin rule [16]. Continuous predictors such as the laboratory tests and age were not converted into categorical predictors.

**Figure 2.**
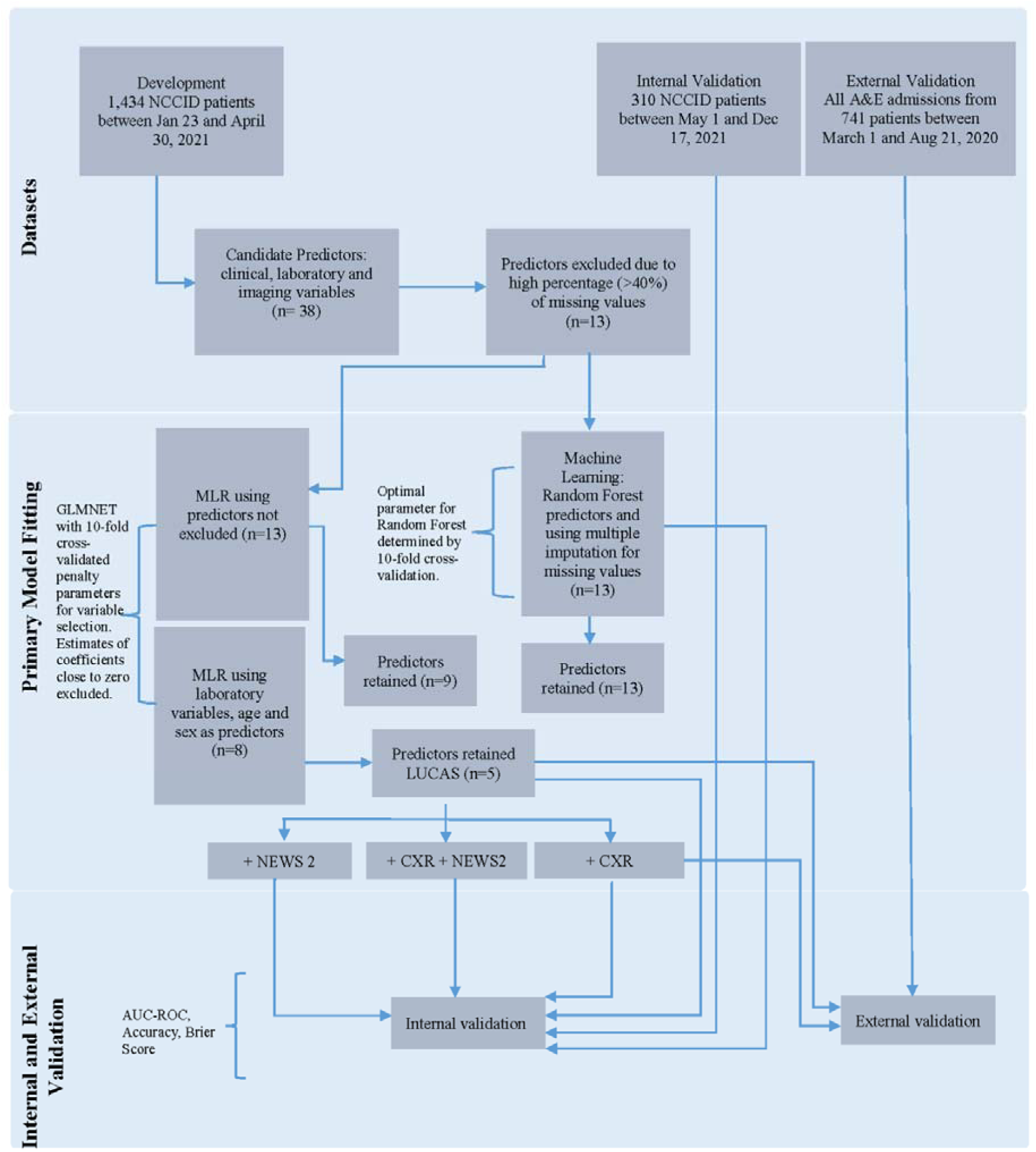
Model development, internal and external validation workflow. AUC-ROC: area under the receiver operating characteristic curve; GLMNET: Lasso and Elastic-Net Regularized Generalized Linear Models; MLR: Multivariable Logistic regression; LUCAS: Lymphocyte, Urea, CRP, Age, Sex; CXR: Chest X-ray; NEWS2: National Early Warning Score

An initial multivariable logistic regression (MLR) model started with 13 predictors with less than 40% missing values (Lymphocyte, Urea, CRP, Age, Sex, WCC, Creatinine, Platelets, Diastolic BP, Systolic BP, Temperature, Heart rate, PaO_2_) were used to fit a standard logistic model. Elastic Net (GLMNET) [17], weighted combination of Least Absolute Shrinkage and Selection Operator (LASSO) and ridge regression were used to select predictors of importance. The optimal value of the shrinkage estimator for LASSO and the weight of GLMNET were chosen using a 10-fold cross validation using the R-package caret [18]. Additionally, the performance of the MLR model was compared with the performance of random forest machine learning approach [19].

Another MLR model took a more pragmatic approach in predictor selection that was based on the predictor’s clinical relevance in monitoring COVID-19 infection. This model started with 8 predictors (Lymphocyte, Urea, CRP, Age, Sex, WCC, Creatinine, Platelets) that were retained after GLMNET, LASSO and ridge regression became the final model. In addition, the model’s performance was evaluated if the inclusion of either NEWS2 or CXR or both improved this model’s performance.

Each model’s performance was assessed using Area Under the Receiving Operator Characteristics Curve (AUC-ROC) with a 95% confidence interval. The internal and external validation of each model were assessed with bootstrap resampling. The predictor selection and entire modelling process were repeated in 1000 bootstrap samples to estimate the realistic predictive measures for future patient cohorts. An AUC-ROC of 0.5 indicated a prediction based on random chance, 0.7 to 0.8 was considered acceptable, above 0.8 demonstrated excellent and a value of 1.0 showed a perfect prediction [20]. We also evaluated the goodness of fit using the Brier score [21] which is a measure to quantify the closeness of the probabilistic predictions to the binary ground-truth class labels. The score varies between 0 and 1, with the lower score indicating superior performance. Finally, the accuracy of the models was evaluated using the standard cut-off value of 0.5.

The internal validation included the NEWS2 score; however, NEWS2 score was not available for the external validation dataset. The statistical package R (3.5.3) was used to perform all statistical analysis and the methods employed in the model development, validation workflow and primary model fitting followed by both internal and external validation.

### Data protection / Ethics

De-identified and pseudo-anonymised patient data were obtained from data sets which were approved by the ethics committee as part of the existing Cardiac MRI Database NHS REC IRAS Ref: 222349 and University of Brighton REC (8011).

### Data availability

The data that support the findings of this study are available from the NCCID [12] and another NHS site; but restrictions apply to the availability of these data sets, which were used under license for the current study, and are not publicly available.

### Code availability

The Simple Logistic Regression based calculator is freely available at https://mdscore.net/

## Results

Overall, the NCCID database comprised of 3818 patients (23/01/2020 - 31/12/2020). The number of patients that met the inclusion criteria and missingness pattern were 1434 and 310 patients for the development and internal validation data sets, respectively (**Figure 1**). All available data on positive patients were used in the external validation data set and divided between 242 fatal (mortality) and 499 non-fatal (no mortality) cases. The development dataset included 38 data points (continuous and categorical) that followed the removal of date data and ≥40% missingness. Summary statistics of median (IQR) for numeric variables and proportions for categorical variable are shown and compared to the internal validation and external validation data sets (**Table 1)**.

### Model Development

From the NCCID 38 possible predictors we focused on 13 predictor variables based on clinical reasoning that related to demographics, clinical observations, laboratory parameters recorded at the time of admission and did not have more than 40% missing data (**Figure 2**). The variable importance plot indicates the relative importance of these predictors (**Figure 3)**. The GLMNET model selection criteria optimally chose predictors based on 10-fold cross validation that created the MLR model and had an AUC-ROC of 0.759 (**Table 2)**. In contrast, the random forest (RF) model used all 13 predictors which increased the complexity of the prediction and achieved a slightly higher AUC-ROC value of 0.806. Thus, the simplicity and ease of fitting our MLR model makes it a robust score which can be easily adaptable to new training data.

**Table 2.** Best Fit Model by Lasso based sequential variable selection.

|  | Predictors |  |  |  |  |  |  |  |  |  |  |  |  |  | Model Performance |  |
| --- | --- | --- | --- | --- | --- | --- | --- | --- | --- | --- | --- | --- | --- | --- | --- | --- |
| Model<br>Process /<br>(number of<br>predictors<br>in initial<br>model) | Lymphocyte | Urea | CRP | Age | Sex | WCC | Creatinine | Platelets | Diastolic BP | Systolic BP | Temperature | Heart Rate | PaO<sub>2</sub> | AUC<br>ROC | Sensitivity | Specificity |
| MLR<br>(n = 8) | Y | Y | Y | Y | Y | N | Y | N | N | N | Y | N | Y | 0.759 | 0.935 | 0.295 |
| Random<br>Forest<br>(n = 13) | Y | Y | Y | Y | Y | Y | Y | Y | Y | Y | Y | Y | Y | 0.860 | 0.909 | 0.573 |
| LUCAS<br>(n = 5) | Y | Y | Y | Y | Y | N | N | N | x | x | x | x | x | 0.765 | 0.927 | 0.315 |

CRP: C-Reactive Protein; WCC: White Cell Count; ROC: receiver operator curve; BP: Blood Pressure; PaO<sub>2</sub>: Partial Oxygen Saturation; Y = retained in the model; N = excluded from the model; x = not used in the model

**Figure 3.**
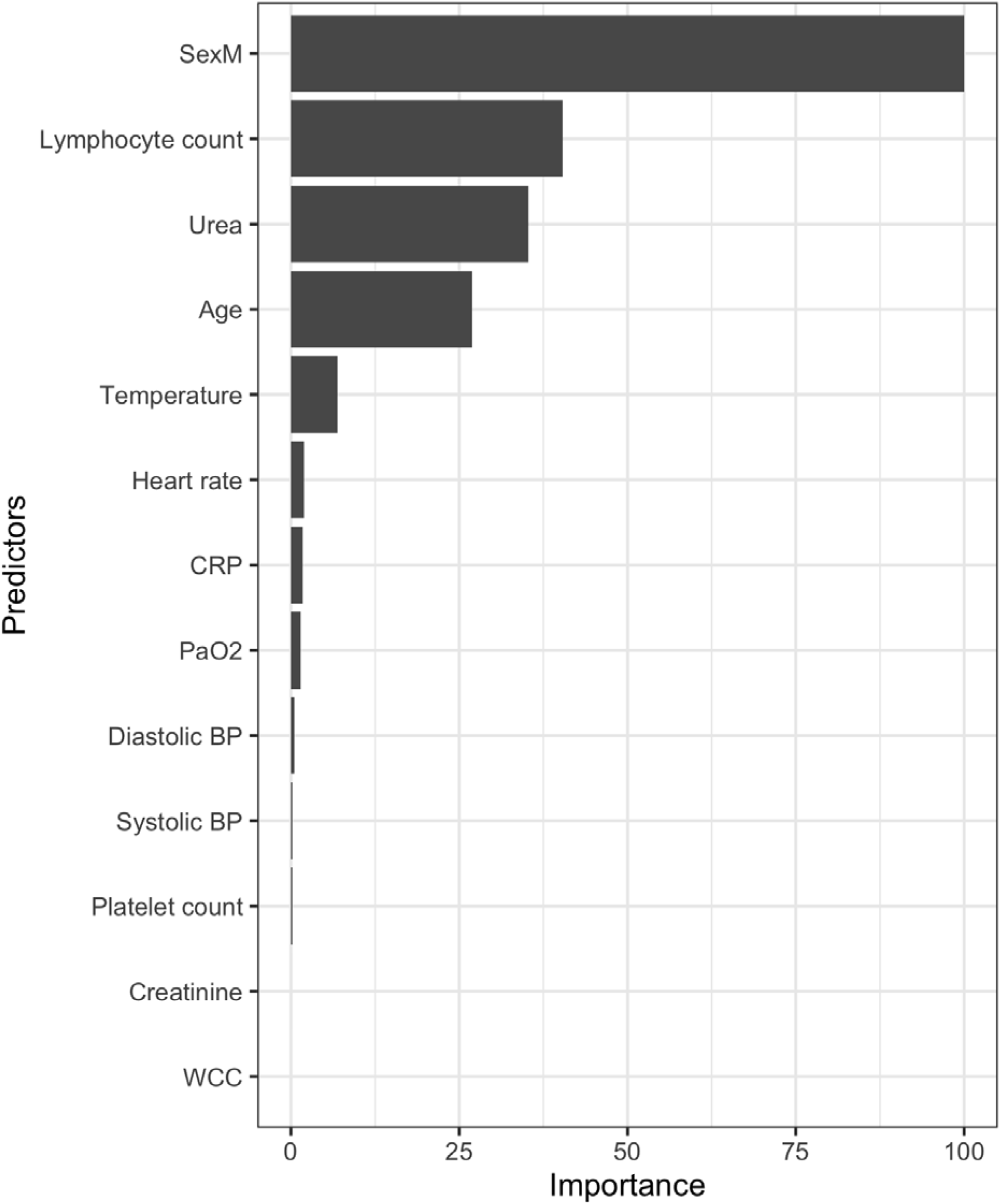
GLMNET (Lasso/Ridge) Variable importance plot with variables having less than 40% missing values in the database. All patients are confirmed positive for SARS-CoV-2 by RT-PCR. The plot shows the importance of variables in building the development predictive model. BP: blood pressure; PaO_2_: partial pressure of O_2_; WCC: white cell count; CRP: C-Reactive Protein.

The rapid blood parameter measurements, sex and age recorded at the time of admission and they parameterized our model. The relative difference of the laboratory parameters along with age, between patients with fatal and non-fatal outcome are presented in **Figure 4**.

**Figure 4.**
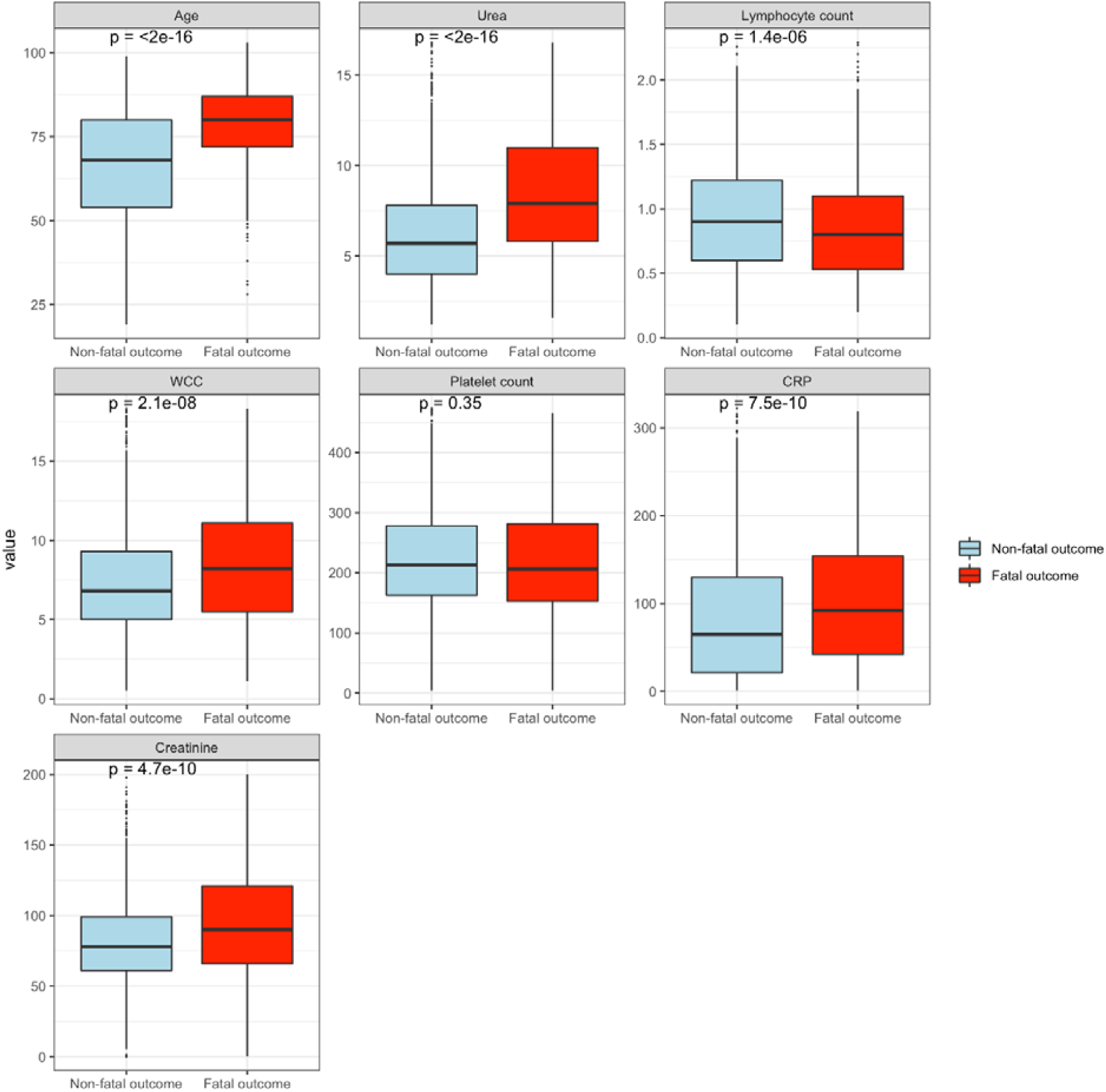
Boxplots showing median and 1st/3rd quartile of individual blood variables parameters plus age categorized by patient outcome (fatal; red box or non-fatal; blue box). All NCCID patients tested positive by the RT-PCR test for SARS-CoV-2. WCC: White blood count; CRP: C-Reactive Protein. The *p*-values are tests of equality of population using the Wilcoxon rank sum test, where *p* < 0.05 implies statistically significant difference between the populations.

Starting the MLR model with these eight predictors, namely, lymphocyte, urea, CRP, age, sex, WCC, creatinine, platelets, and using GLMNET for variable selection, a final model of 5 predictors were retained with AUC-ROC 0.765 (**Figure 5**). These predictors: **L**ymphocyte count, **U**rea, **C**RP, **A**ge and **S**ex (LUCAS) became the final model with optimal accuracy results compared to the MLR and random forest models with different number and set of predictors included (**Table 2**). From these results, the model can accurately predict 91% of the true fatal outcomes and 36% of the true non-fatal outcomes.

**Figure 5.**
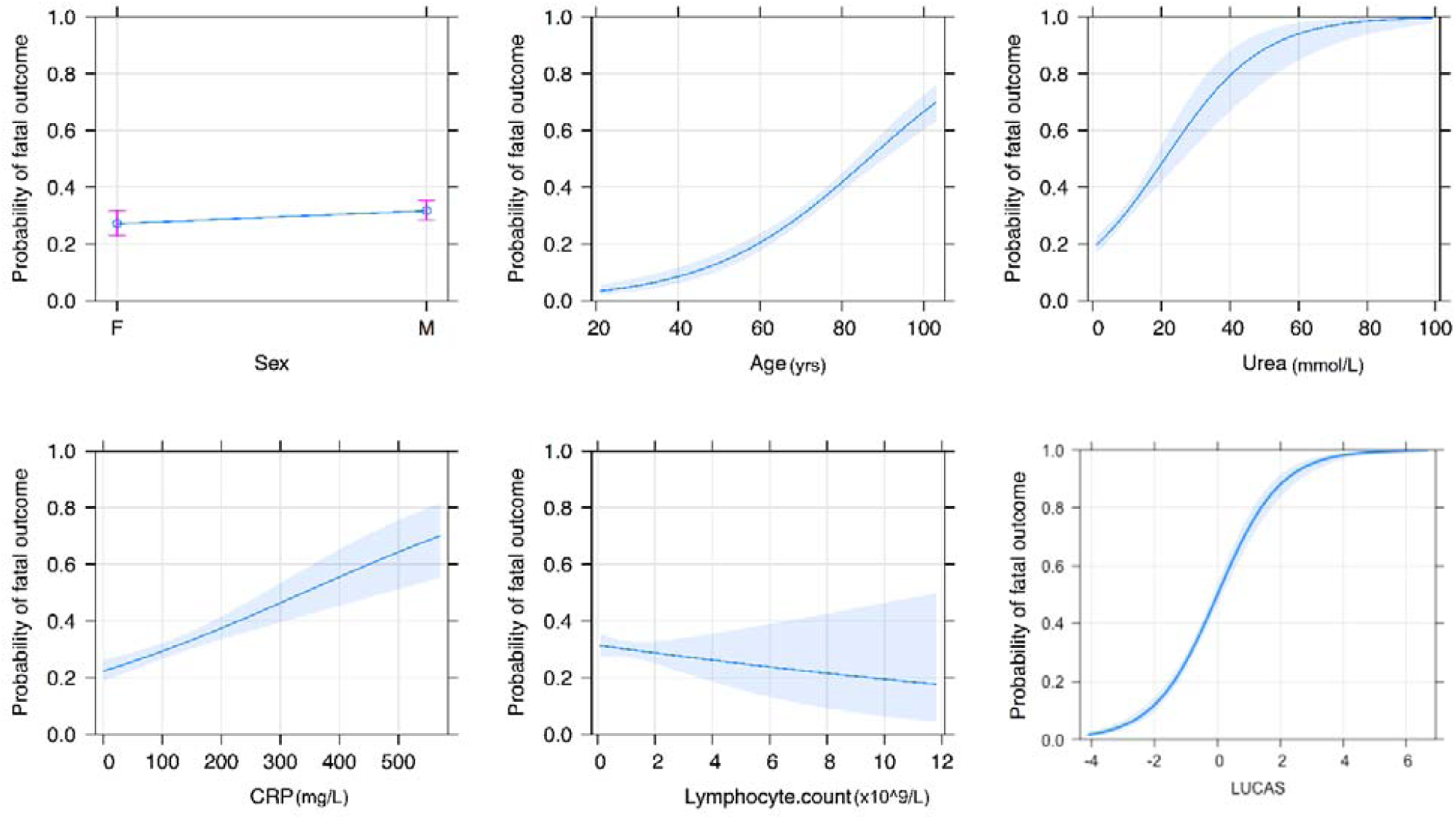
Predictor plots; Multivariate association between predictors (Age, Sex, Urea, CRP and Lymphocyte count) and probability of fatal outcome.

The first step taken to stratify the outcomes from each predictor involved effect plots along with the estimated linear predictor (linear combination of the predictors with the estimated coefficients of MLR) (**Figure 5**). Each sub-plot in **Figure 5** indicates how the individual predictor is related to the probability of fatal outcome. The binary predictor of sex shows an increase in probability of fatal outcome for males. For the continuous predictor of age, urea and CRP, we can see a clear increase in the probability of fatal outcome corresponding to the increase in value of these predictors. We can also observe that in the presence of other predictors, lymphocyte count is a weak predictor as there is a slight decrease in the probability of fatal outcome with the increase in lymphocyte count. We can also notice that the error band around the mean effects is quite wide on the higher values of CRP and lymphocyte count since there are only a few observations in the higher range of these two predictors. Finally, we can clearly see that the linear combination given by LUCAS has a steep slope with a very narrow error bar giving us a very sensitive and accurate mortality score.

Along with the exact values of the probability of fatal outcome based on the five predictors, we propose a risk stratification (**Table 3**) based on these probabilities which allows a simplified interpretation of mortality to low, medium, high and very high risk of a fatal outcome.

**Table 3.** Risk stratification of LUCAS

| **Risk Level** | **Probability**<br>**of Fatal**<br>**Outcome**<br>**cut-off** | **Interpretation**<br>**(60-day**<br>**mortality)** | **Proportion in**<br>**Derivation Set**<br>***n* (%)** | **Proportion in**<br>**Internal Validation**<br>**Set *n* (%)** | **Proportion in**<br>**External Validation**<br>**Set *n* (%)** |
| --- | --- | --- | --- | --- | --- |
| **Low** | < 0.235 | 0.8 - 1.2 % | 574 (40) | 127 (41) | 201 (27) |
| **Moderate** | 0.235 - 0.465 | 33 - 40 % | 516 (36) | 112 (36) | 333 (45) |
| **High** | 0.465 - 0.7 | 53 - 60 % | 258 (18) | 59 (19) | 170 (23) |
| **Very High** | >0.7 | >65 % | 86 (6) | 12 (4) | 37 (5) |

Though the percentage of missing predictors in our reduced models were negligible, sensitivity analysis was performed using complete case data and as expected the model performance did not change. We also note that, the simple logistic regression was preferred over machine learning approaches such as Random Forest starting from same set of predictors, based on simplicity and their performance in the validation set.

### Model Validation

We have performed validation on an internal validation (NCCID Data between 1^st^ May 2020 and 7^th^ December 2020) cohort and external validation cohort consisting of data from a separate A+E site (n = 1012; between 1^st^ March 2020 and 21^st^ August 2020) with a small overlap with the available predictors from the derivation and internal validation cohort. However, all five predictors from the model were present (LUCAS: **L**ymphocyte count, **U**rea, **C**RP, **A**ge and **S**ex).

The median age of patients from the internal validation dataset was 74 years (interquartile range; IQR 62-84), which was not significantly different from the external validation dataset with a median of 76 years (IQR 61-84). The proportion of male patients was similar in both groups; 58% male in the internal validation set, and 55% male in external validation set (**Table 1**).

The performance of the LUCAS calculator on the internal validation cohort was very good; AUC-ROC 0.744 (CI: 0.673 - 0.808) with an accuracy 0.796 at the standard cut-off of 0.50 for the probability of a SARS CoV-2 positive patient dying within 60 days of a positive test (**Table 4**). Use of the available NEWS2 data in combination with LUCAS had little effect on accuracy (AUC-ROC 0.747, CI: 0.668 - 0.821). In contrast, inclusion of CXR data in combination with LUCAS led to a slight increase in AUC-ROC to 0.770 (CI: 0.695 - 0.836). The choice to use the CXR outcome (normal vs abnormal) was therefore included in the simple LUCAS calculator as an optional extra predictor.

**Table 4.** Internal and External Validation results.

| **Model** | **AUC-ROC** | **95% confidence interval (upper to lower)** | **Accuracy at 0.50 cut-off** | **Brier Score** |
| --- | --- | --- | --- | --- |
| **Development** |  |  |  |  |
| **(Data between 23/01/2020 - 30/04/2020) $n = 1434$** |  |  |  |  |
| LUCAS | 0.765 | 0.738 - 0.790 | 0.726 | 0.179 |
| CXR | 0.550 | 0.527 - 0.572 | 0.451 | 0.219 |
| NEWS2 | 0.591 | 0.555 - 0.629 | 0.691 | 0.211 |
| LUCAS + CXR | 0.774 | 0.748 - 0.802 | 0.732 | 0.175 |
| LUCAS + NEWS2 | 0.771 | 0.742 - 0.799 | 0.751 | 0.173 |
| LUCAS + NEWS2 + CXR | 0.779 | 0.746 - 0.808 | 0.742 | 0.171 |
| **Internal Validation** |  |  |  |  |
| **(Data between 01/05/2020 - 07/12/2020) $n = 310$** |  |  |  |  |
| LUCAS | 0.744 | 0.673 - 0.808 | 0.796 | 0.156 |
| CXR | 0.596 | 0.528 - 0.653 | 0.447 | 0.165 |
| NEWS2 | 0.655 | 0.570 - 0.736 | 0.711 | 0.159 |
| LUCAS + CXR | 0.770 | 0.695 - 0.836 | 0.794 | 0.152 |
| LUCAS + NEWS2 | 0.747 | 0.668 - 0.821 | 0.778 | 0.154 |
| LUCAS + NEWS2 + CXR | 0.757 | 0.670 - 0.831 | 0.759 | 0.158 |
| **External Validation** |  |  |  |  |
| **(Data between 01/03/2020 - 21/08/2020) $n = 741$** |  |  |  |  |
| LUCAS | 0.752 | 0.713 - 0.790 | 0.706 | 0.187 |
| LUCAS + CXR | 0.791 | 0.746 - 0.833 | 0.714 | 0.165 |
| CXR | 0.517 | 0.476 - 0.559 | 0.448 | 0.179 |

LUCAS: Lymphocytes, Urea, CRP, Age, Sex model; CXR: Chest X-ray severity score; NEWS2: National Early Warning Score 2

LUCAS predictor outperformed in external validation by 0.752 (CI: 0.713-0.790) AUC-ROC value contrary to the internal dataset (0.744 CI: 0.673 - 0.808). This can justify the robustness and generalization of the calculator since the variation between the two datasets is not high (0.046, robust) and the model outperforms in the external validation (generalization). Including CXR information improved the prediction performance of AUC-ROC to 0.791 (CI: 0.746 - 0.833). Note that, the NEWS2 score was not available for the external validation cohort, so this result could not be externally validated.

## Discussion

We have developed and validated a simplified, fast track mortality calculator based on three rapid and routine simple predictors plus age and sex, which can be optionally used in combination with CXR results. The LUCAS calculator is freely available, relies on objective measurements only, and has been both internally and externally validated. The use of LUCAS calculator is for use to aid triage on patient admission to A&E following a positive SARS CoV-2 RT PCR test. The robustness and generalized results in the validation process classify the tool as an excellent candidate for risk management of the mortality level in a 60-day survival interval of adult SAR CoV-2 positive patients. The generalisation of LUCAS was justified since the calculator delivered higher accuracy of the external validation compared with the internal validation set. In addition, the incorporation of CXR results as normal versus abnormal improved the prediction performance.

The NCCID dataset collected 38 clinical data points for each patient that served as candidate predictors. These were categorised into demographics, risk factors, past medical history, medications, clinical observations, chest-imaging data, and laboratory parameters measured on admission. All laboratory tests were measured before SARS CoV-2 RT-PCR test and mortality date, which ensured these potential predictors were blinded to the outcome. The SARS CoV-2 swab and RT-PCR results established the final COVID-19 status. Moreover, thorough study of other included measures in the event of deterioration, such as the NEWS2 score, was also evaluated. As a result, a comprehensive study and evaluation of all the possible blood markers and clinical patient information was taken into consideration. This study shows that a simple and objective tool can risk stratify SARS CoV-2 positive patients within one hour after hospital admission. The primary objective showed that rapid and routine laboratory blood tests and chest imaging data added predictive value beyond the RT-PCR test and clinical observations with high AUC-ROC.

### Comparison with other studies

There have been a large number of prognostic tools published, most notably the 4C Mortality Score [22] and QCOVID [23] which were based on very large cohort datasets and includes a large number of predictors in their algorithms. Our study is the first to combine a minimum number of blood results along with CXR data, so as to generate a simplified calculator based on as few objective predictors as possible.

The 4C Mortality Score includes a large number of parameters including PMH, demographics and blood measurables, which results a high AUC ROC of 0.79. However, gaining an accurate past medical history during triage is not always practical, and the 4C calculator was not externally validated, and its use has been limited due to its complexity. Our aim was to use the minimum number of predictors without losing accuracy, which was achieved, resulting in LUCAS which exhibits a similar level of prediction as the more complex and detailed 4C algorithm. The primary QCOVID score was developed as a risk prediction algorithm to estimate hospital admission and mortality outcomes, which also included a large number of predictors, including PMH. More importantly the authors clearly state that their score is “not intended for use supporting or informing clinical decision-making.”

Numerous prediction models have been developed to aid triage and research into COVID-19 disease severity. While there has been a great deal of useful insight into the disease gained from these studies and prognostic tools, there is a range of outcomes mostly due to some having a high risk of bias, lack of transparency and lack of internal [24] and or external validation [22, 25]. Our study improves on these issues by conforming to PROBAST and being both internally validated from the same large NCCID dataset, and externally validated in a smaller, separate hospital database. In addition, many studies require past medical history, or base the prediction solely mainly on the underlying health conditions of the patient [25, 26]. These data may be difficult to assess accurately on admission to hospital, and may mislead should the patient have undiagnosed conditions. For this reason, we focused our model on measurables taken routinely on admission.

Inclusion of CXR data is optional on the online LUCAS calculator, and based on simple outcome of normal/abnormal image results. The ability to include CXR results is not widely used in other prediction-modelling calculators and has been included in one other study [25] along with ten other parameters (symptoms, past medical history and measurables). Our focus however was to form a simplified model on blood results and with the option of CXR images, which we have achieved.

All of the predictors used in LUCAS have individually been indicated as important in COVID-19 severity and mortality, although our study is the first to use only these predictors in a prognostic score. Leukocytes, Urea and CRP are recognised as key measurable predictors of severity of SARS CoV-2 infection, and age and sex are well-known predictors of mortality [22, 23, 27].

Throughout the study, we have carefully considered the risk of bias that is inherent in retrospective studies. By conducting both internal and external validation the study here indicates a robust model, with reduced bias since only patients testing positive for SARS CoV-2 were included in the development of the LUCAS algorithm. The size of the external validation set was smaller than the development set allowing us to check for discrimination of population size, and the results indicate that the LUCAS calculator can predict form small cohorts as well as it can from larger size populations. The patient data was collected at an early stage of the pandemic, when treatments differed to later in the year, which would affect the death rate in hospital, and our results do not account for non-hospital death that may occur outside of the hospital or in the 60-day window following diagnosis. Over time, any algorithm of mortality will change due to improvements in therapies as well as the use of vaccination which will change the profile of those at risk of COVID-19 related death.

In conclusion, there are major strengths of this study including the analytical approach taken, the comparison to NEWS2 score, the inclusion of CXR data and the resulting algorithm is both internally and externally validated. The CXR data contributes an improvement in the prediction of the mortality of the patient, however the results used in the LUCAS calculator is basic. Future development of this work will be to extract pixel to pixel information based on CXR medical image analysis using deep learning or machine learning methods. Thus, as feature work we will combine LUCAS with ML and DL models to deliver the option of CXR imaging analysis when this is available.

## Data Availability

https://mdscore.net

## Footnotes

### Contributors

SW, LM are joint last authors. LM is corresponding author for this article.

### Contributor Roles

Contributor Roles: Conceptualisation: SR, AS, AB, MM, SW, LM; Data Curation: SR, AB, BV; Formal Analysis: SR; Funding Acquisition: SR, AS, LM; Investigation: SR; Methodology: SR, AS, AB, MM, BV; Project Administration: SW, JF; Resources: BV, AS; Software: SR, AB, BV; Supervision: SW; Validation: SR, AB, MM, BV; Visualisation: SR, SW, LM; Writing Original Draft: SW, LM, SR; Writing – review and editing: SW, LM, SR, AS, MM, AB, BV

### Funding

EPSRC Impact Acceleration account fund EP/R511705/1; AS is funded by a Wellcome Trust fellowship 205188/Z/16/Z; University of Brighton COVID-19 Research Urgency Fund

### Competing interests

All researchers declare no competing interests.

## Acknowledgements

The researchers would like to thank the biomedical scientists at NHS Greater Glasgow and Clyde hospitals and Portsmouth Hospital NHS Trust for their advice.

## Notes

### Competing Interest Statement

The authors have declared no competing interest.

