## Supplementary Table and Figures for "LUCAS: A highly accurate yet simple risk calculator that predicts survival of COVID-19 patients using rapid routine tests"

**Supplementary Table 1.** Origins of Dataset: development and internal validations (NCCID NHS Trust Summary) <https://nhsx.github.io/covid-chest-imaging-database/>

|  | Total | Male | Female | Dead | Alive | Prop. dead |
| --- | --- | --- | --- | --- | --- | --- |
| Ashford and St Peters Hospitals NHS Foundation Trust | 139 | 59 | 80 | 133 | 6 | 0.96 |
| Betsi Cadwaladr University Health Board | 114 | 67 | 47 | 41 | 73 | 0.36 |
| Brighton and Sussex University Hospitals NHS Trust | 248 | 136 | 112 | 99 | 149 | 0.40 |
| Cambridge University Hospitals NHS Foundation Trust | 0 | 0 | 0 | 0 | 0 | NA |
| Cwm Taf Morgannwg University Health Board | 90 | 51 | 39 | 24 | 66 | 0.27 |
| George Eliot Hospital NHS Trust | 665 | 394 | 271 | 234 | 431 | 0.35 |
| Hampshire Hospitals NHS Foundation Trust | 205 | 117 | 88 | 50 | 155 | 0.24 |
| Imperial College Healthcare NHS Trust | 1708 | 1042 | 666 | 456 | 1252 | 0.27 |
| Liverpool Heart and Chest NHS Foundation Trust | 61 | 53 | 8 | 26 | 35 | 0.43 |
| London North West University Healthcare NHS Trust | 420 | 227 | 193 | 149 | 271 | 0.35 |
| Norfolk and Norwich University Hospitals NHS Foundation Trust | 692 | 384 | 308 | 238 | 454 | 0.34 |
| Oxford University Hospitals NHS Foundation Trust | 24 | 15 | 9 | 14 | 10 | 0.58 |
| Royal Cornwall Hospitals NHS Trust | 759 | 523 | 236 | 300 | 459 | 0.40 |
| Royal Surrey NHS Foundation Trust | 1 | 0 | 1 | 1 | 0 | 1.00 |
| Royal United Hospitals Bath NHS Foundation Trust | 1533 | 987 | 546 | 417 | 1116 | 0.27 |
| Sandwell and West Birmingham Hospitals NHS Trust | 0 | 0 | 0 | 0 | 0 | NaN |
| Sheffield Childrens NHS Foundation Trust | 1 | 1 | 0 | 0 | 1 | 0.00 |
| Taunton and Somerset NHS Foundation Trust | 56 | 44 | 12 | 16 | 40 | 0.29 |
| West Suffolk NHS Foundation Trust | 515 | 330 | 185 | 195 | 320 | 0.38 |

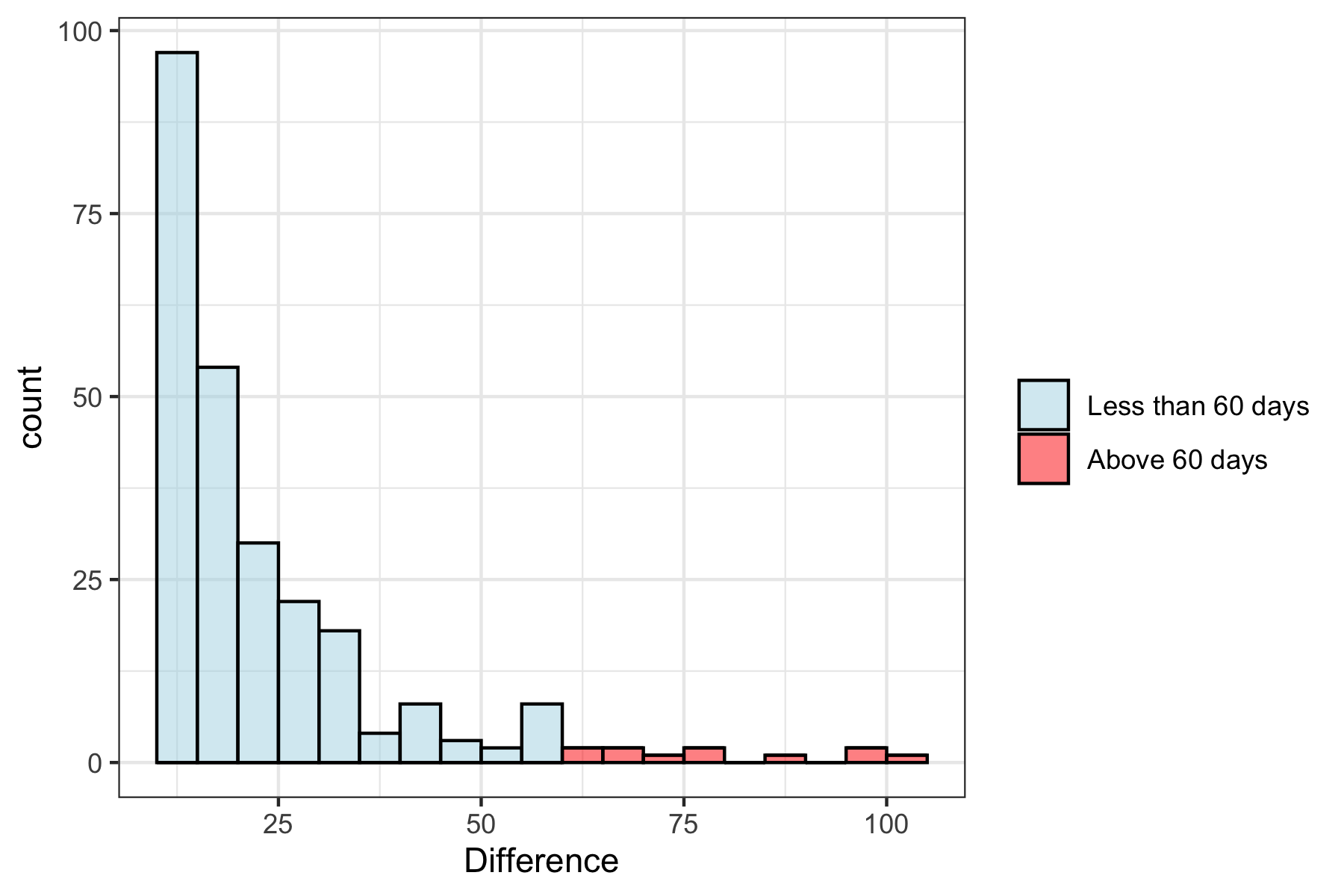

**Supplementary Figure 1.** Histogram showing the number of patients (count) from time of admission to hospital (Internal development cohort 23/01/20 - 30/04/20) with a positive SARS CoV-2 test by RT-PCR and date of fatal outcome. The study conducted included only patients with a fatal outcome within 60 days of hospital admission.

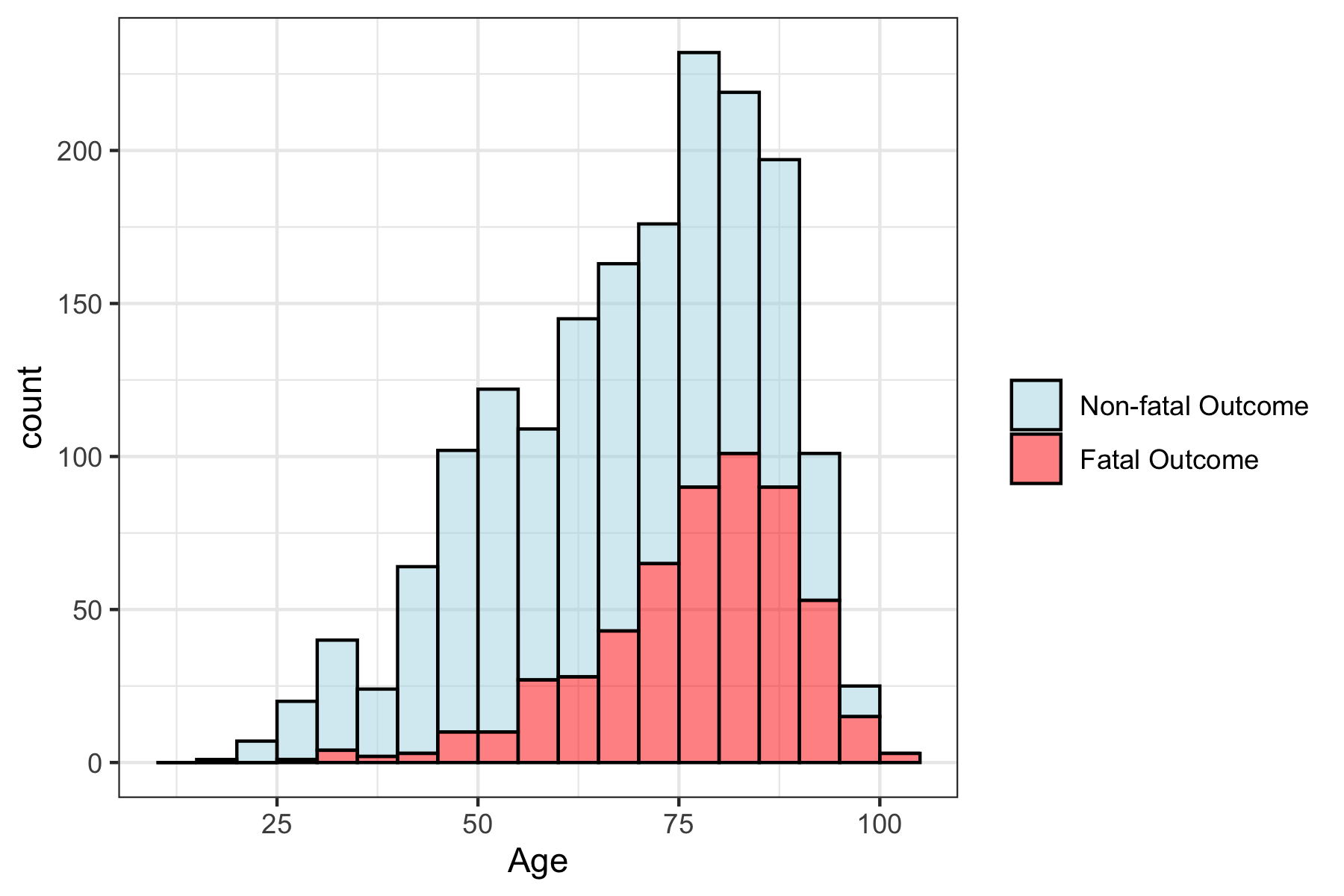

**Supplementary Figure 2.** Characteristics of development dataset. Distribution of age of patients admitted to A+E, with fatal and non-fatal outcome distribution defined within 60 days of a positive test for SARS CoV-2 by RT-PCR.

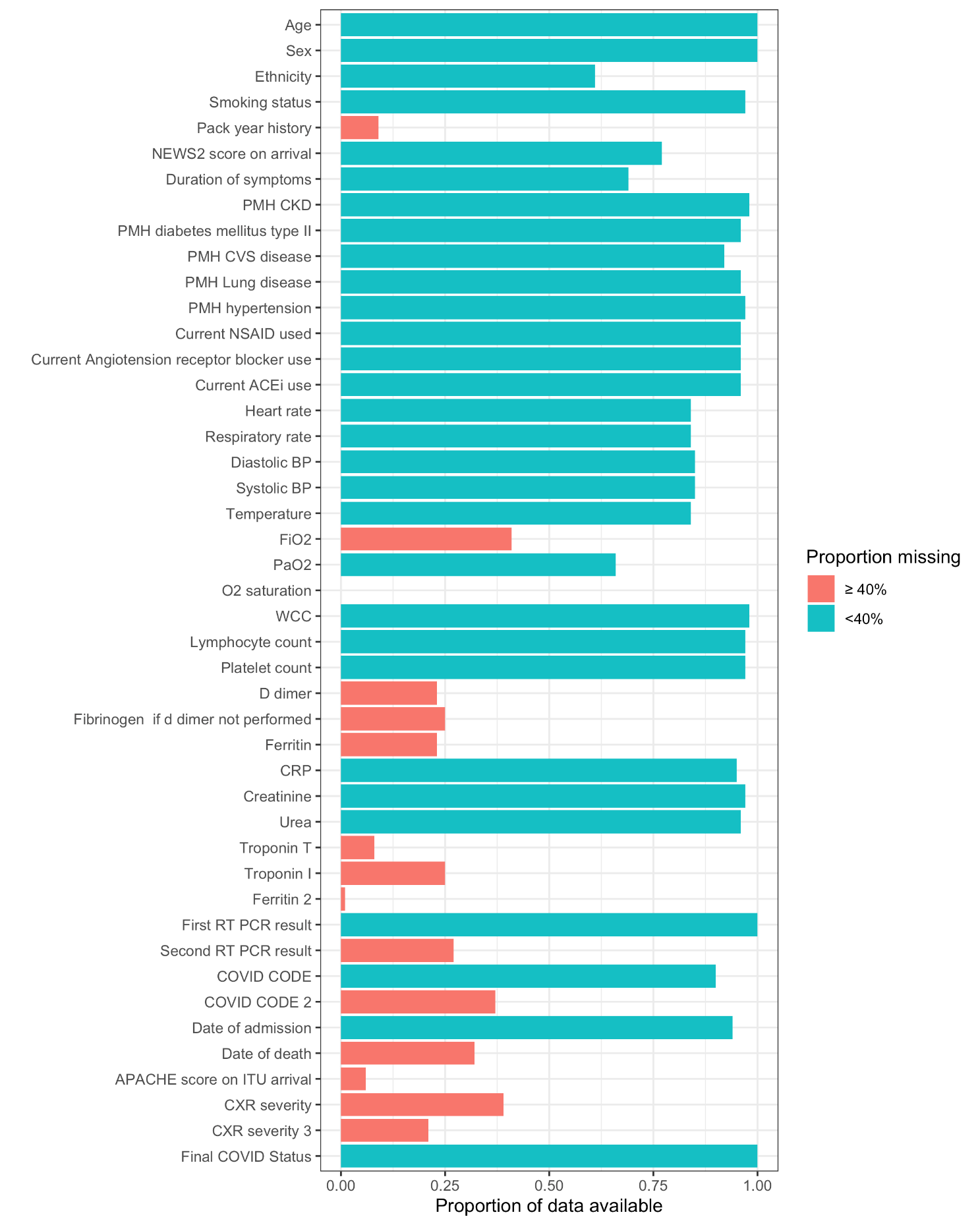

**Supplementary Figure 3.** Density map of data points from NCCID data points to determine predictors to be used in both Development and Internal Validation dataset. Data above the 60% threshold were used in the analysis.

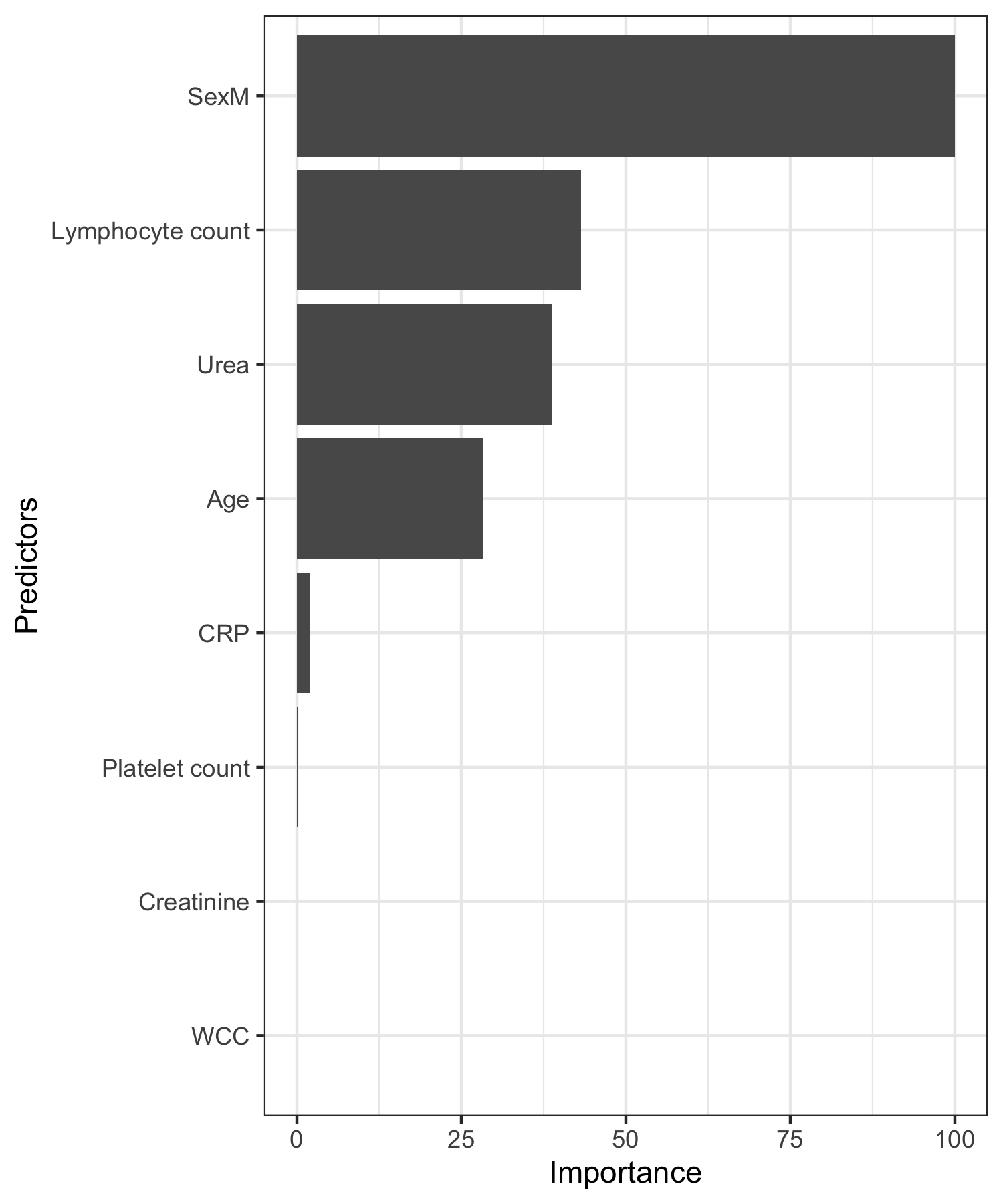

**Supplementary Figure 4.** Variable importance plot with variables having less than 40% missing values in the data set starting from only blood variables (plus age and sex)
